# Innovative quantitative PCR assays for the assessment of HIV-associated cryptococcal meningoencephalitis in Sub-Saharan Africa

**DOI:** 10.1101/2023.08.24.23294467

**Authors:** Tshepiso Mbangiwa, Aude Sturny-Leclère, Kwana Lechiile, Cheusisime Kajanga, Timothée Boyer-Chammard, Jennifer C. Hoving, Tshepo Leeme, Melanie Moyo, Nabila Youssouf, David S. Lawrence, Henry Mwandumba, Mosepele Mosepele, Thomas S Harrison, Joseph N Jarvis, Olivier Lortholary, Alexandre Alanio, The Ambition Study Group

## Abstract

**Background:** Cryptococcal meningitis (CM) accounts for about 10-20% of AIDS-defining illnesses with a 10-week mortality rate of 25-50%. Fungal load assessed by colony-forming unit (CFU) counts is used as a prognostic marker and to monitor response to treatment in research studies. PCR-based assessment of fungal load could be more rapid and less labor-intensive.

**Methods:** We developed and validated species-specific qPCR assays based on DNA amplification of a Quorum Sensing Protein 1 (*QSP1*); *QSP1*A, *QSP1* B/C, and *QSP1* D that are specific to *C. neoformans*, *C. deneoformans* and *C. gattii* species, respectively, and a pan-*Cryptococcus* assay based on a multicopy *28S rRNA* gene. We tested these assays for species identification (QSP1) and quantification (QSP1 ans 28S) on cerebrospinal fluid (CSF) of 209 CM patients at baseline (Day 0) and during anti-fungal therapy (Day 7 and Day 14), from the AMBITION-cm trial in Botswana and Malawi (2018-2021).

**Findings:** When compared to quantitative cryptococcal culture (QCC) as the reference, the sensitivity of the *28S rRNA* and *QSP1* assays were 98.2% [95% CI: 95.1-99.5] and 90.4% [95% CI: 85.2-94.0] respectively in cerebrospinal fluid (CSF) at Day 0. Quantification of the fungal load with *QSP1* and *28S rRNA* qPCR correlated with QCC (R^2^=0.73, R^2^=0.78, respectively). Both Botswana and Malawi had a predominant *C. neoformans* prevalence of 67% [95% CI: 55, 75] and 68% [95% CI: 57, 73], respectively and lower *C. gattii* rates of 21% [95% CI: 14, 31] and 8% [95% CI: 4, 14], respectively. We identified 10 patients that, after 14 days of treatment, harboured viable but non-culturable yeasts based on *QSP1* RNA detection (without any positive CFU in CSF culture).

**Interpretation:** *QSP1* and *28S rRNA* assays are useful in identifying *Cryptococcus* species. qPCR results correlated well with baseline QCC and showed a similar decline in fungal load during induction therapy. These assays have a quick turnaround time and could be used in place of QCC to determine fungal load clearance. The clinical implications of the detection of possibly viable but non-culturable cells in CSF during induction therapy remain unclear.

**Funding:** The AMBITION-cm clinical trial which was funded by the European and Developing Countries Clinical Trials Partnership; Swedish International Development Cooperation Agency; Wellcome Trust / Medical Research Council (UK) / UKAID Joint Global Health Trials and National Institute for Health Research (UK).

## Background

HIV-associated cryptococcal meningitis is the second leading cause of all AIDS-related mortality ^1^. An estimated 152,000 cases of cryptococcal meningitis occur each year, leading to approximately 112,000 deaths ^2^. Even with the best available antifungal regimens, ten-week mortality is 25-30% ^3,4^.

*Cryptococcus neoformans* and *Cryptococcus gattii* species complexes are responsible for cryptococcosis in humans, with the former being the predominant causative organism with elevated morbidity and mortality in immunocompromised individuals and the latter predominant in immunocompetent persons, at least in certain areas.^5^ *C. neoformans* species complex can be characterized into serotype A (*C. neoformans*), serotype D (*C. deneoformans)* and A-D hybrids, while *C. gattii* species complexes are subdivided into two, serotype B and serotype C including *C. gattii*, *C. bacillisporus*, *C. deuterogattii*, *C. tetragattii*, *C. decagattii* ^6^. qPCR assays have already been developed to discriminate these species on colony DNA ^7–9^, however, these assays are not suitable for diagnosis, following MIQE guidelines ^10^.

Methods used to diagnose cryptococcal meningitis include India ink staining, fungal culture and cryptococcal polysaccharide capsular antigen (CrAg) detection. CrAg detection can be performed with lateral flow assays (LFA) which are easy and quick to perform.^11–14^ However, LFA is primarily qualitative and semi-quantitative but does not correlate with the fungal load under treatment, which could be important for the timely monitoring of patient prognosis. Kinetics of CrAg in patients on treatment cannot be used as a prognostic marker as they do not correlate well with a decline in QCC, CrAg also remains positive even after patients have cleared CFU via QCC, which limits the usefulness of CrAg testing in patient monitoring. ^14,15^ Colony forming units (CFU) counts using quantitative cryptococcal culture (QCC) are performed in research studies to quantify the fungal load of culturable yeasts in the CSF and evaluate early fungicidal activity (EFA)^16,17^. However, QCC is fastidious and time-consuming. Quantitative PCR (qPCR) allows for DNA quantification and reverse transcriptase PCR (RT-qPCR) allows for whole nucleic acid (WNA) amplification and evaluation of viability by detecting mRNA ^18–21^ and is a potential alternative method to monitor patient response to treatment ^20^.

We, therefore, developed and evaluated four qPCR assays to identify *Cryptococcus* species/ species complexes and quantify *Cryptococcus* load directly from the CSF. We then validated our assays using CSF samples from participants enrolled in the AMBITION-cm trial to offer an innovative approach to monitor the fungal load dynamics.

## Methods

### Quantitative PCR assays and primer design

Primers specific to *Cryptococcus neoformans* (serotype A), *Cryptococcus deneoformans* (serotype D) and *Cryptococcus gattii* species complex (serotype B or C) were designed using Primer 3 (http://bioinfo.ut.ee/primer3-0.4.0/) and checked for secondary structures on OligoAnalyzer Tool (https://eu.idtdna.com/calc/analyzer). NCBI primer blast (https://www.ncbi.nlm.nih.gov/tools/primer-blast/) was used to check for *in silico* specificity. Two assays were developed, targeting (i) the single copy gene Quorum sensing protein 1 (*QSP1*) and (ii) the multicopy gene *28S rRNA* gene (Figure 1). *QSP1* is a *Cryptococcus* specific gene with no ortholog and paralogs outside Cryptococcus neoformans and gattii species complexes (https://fungidb.org/fungidb/app/record/gene/CNAG_03012#Taxonomy). The *28S rRNA* gene is known as a repeated gene which is a category of genes that is currently recommended to use to improve the sensitivity of the qPCR assay ^10^. However, the number of copies can vary from an isolate to another ^22^. Therefore, for precise quantification, a unique gene (here *QSP1*) is preferred, when sensitivity of the detection is not an issue. The *28S rRNA* assay was designed to capture the complete diversity within the Cryptococcus neoformans and gattii species complexes in one single assay as designing a specific assay for each recently described species ^6^ is not cost-effective and relevant in routine care.

**Figure 1:**
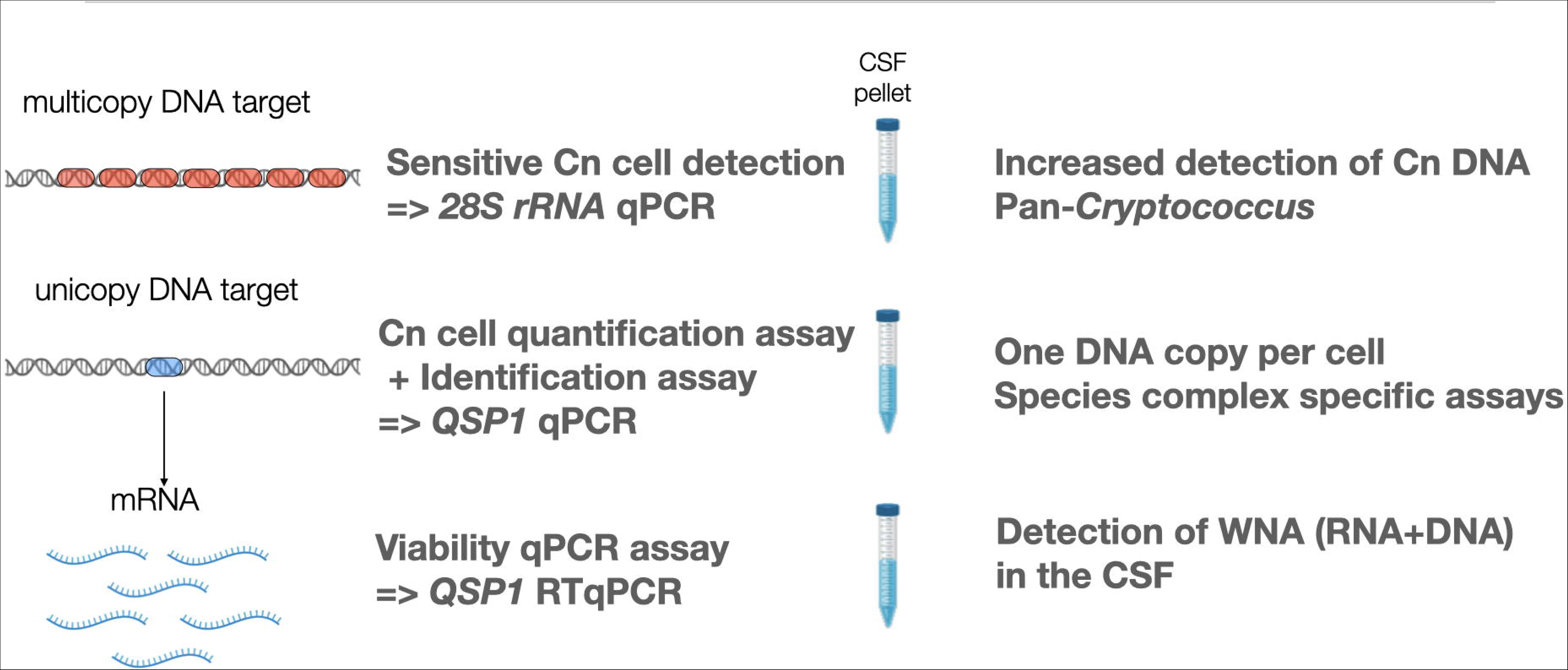
Schematic representation of the different assays used in this study Cryptococcus neoformans (Cn); whole nucleic acids (WNA), CSF (Cerebrospinal fluid), quantitative real time PCR (qPCR), reverse transcriptase quantitative PCR (RT-qPCR).

Three *QSP1* assays specific to serotype A (*QSP1A*), D (*QSP1D*) and B/C (*QSP1B/C*) were designed for identification and precise quantification (one copy corresponding to one cell) (Figure 1). The *28S rRNA* assay was designed to be pan-*Cryptococcus* and to improve sensitivity to detect low fungal loads as present in several copy in the genome. A reverse transcriptase qPCR (RT-qPCR) assay detecting *QSP1* mRNA using *QSP1A* primers was used to validate and check *Cryptococcus* viability in the clinical specimens. Indeed, mRNA is one of the most abundant transcript in the Cryptococcus cells as depicted in FungiDB (https://fungidb.org/fungidb/app/record/gene/CNAG_03012#TranscriptionSummary). As the mRNA transcript is fragile, more than DNA copies, a decreased detection of mRNA as compared to DNA level was considered as proxy of dead yeasts, with only viable cells allowing the production of an increased quantity of mRNA (Supplemental Figure 2F).

### Primer specificity

The primers were tested on the *C. neoformans* reference strain H99 cells seeded in phosphate buffered saline (PBS) (simulated CSF) and in healthy blood samples at different concentrations and stored at −80⁰C for at least 48 hours. This was meant to mimic the storage conditions of the CSF specimens collected during the trial ^3^ before whole nucleic acid extractions were carried out. The assays were tested on all the *C. neoformans* and *C. gattii* species complexes, and a panel of 89 other species (Supplementary table 1) and fourteen human DNA samples to rule out cross-reactivity with human DNA and other fungal species.

### Nucleic acid extractions

For optimization experiments presented in the supplemental material, whole nucleic acids (WNA, representing DNA + RNA) were extracted from PBS seeded with H99 cells using the MagNA Pure 96 Instrument (Roche Diagnostics, Mannheim, Germany) and Viral NA large volume kit (Roche Diagnostics, Mannheim, Germany). Comparison between the DNA blood 1000 and pathogen universal 1000 extraction protocols on the extraction machine was done using PBS spiked with H99 cells (simulated CSF). Different pre-treatment conditions known to improve nucleic acid extractions were tested, including untreated (control), bead beating (BB), adding 50µl of proteinase K (PK) and 10 min incubation at 65°C before pre-extraction, and a combination of BB and PK.

WNA was extracted from stored patient CSF pellet obtained by centrifugation of 1 mL of CSF and freezing. A Pathogen universal 500 protocol was used for extraction and 100ul of WNA was eluted. The RNA process control Roche kit (Roche Diagnostics, Mannheim, Germany) was used as an internal control in RT-qPCR while a DNA internal control kit (DICR-CY5, Diagenode, Seraing, Belgium) was used in qPCR runs, with a defined virus quantity directly added in the sample before extraction, as recommended.^10^

### Quantitative real-time PCR (qPCR) and RT-qPCR

A Light Cycler® 480-II machine (Roche Diagnostics, Mannheim, Germany) was used for all qPCR and RT-qPCR amplification and quantification cycles (Cq) analysis. A Roche Light Cycler 480 probes master kit (Roche Diagnostics, Mannheim, Germany) was used for all qPCR reactions, while a TaqMan™ Fast Virus 1-Step Master Mix kit (Thermo Fisher Scientific Inc, Massachusetts, USA) was used for all RT-qPCR reactions. A 0.5 μM (primer) and 0.2 μM (probe) final concentrations were used for *QSP1*, while 0.4 μM primer and 0.2 μM probe final concentrations were used in the *28S rRNA* assay (Table 1). qPCR reactions were run as following: 95°C 10 min, 50 cycles of 95°C 10 sec and then 58°C 30 sec, and 40°C 30 sec, with an additional initial step at 50°C 5 min for RT-qPCR. All Cq values over 40 were considered negative. All qPCR assays were performed blindly to quantitative cryptococcal culture (QCC) results and all clinical information.

**Table 1:**
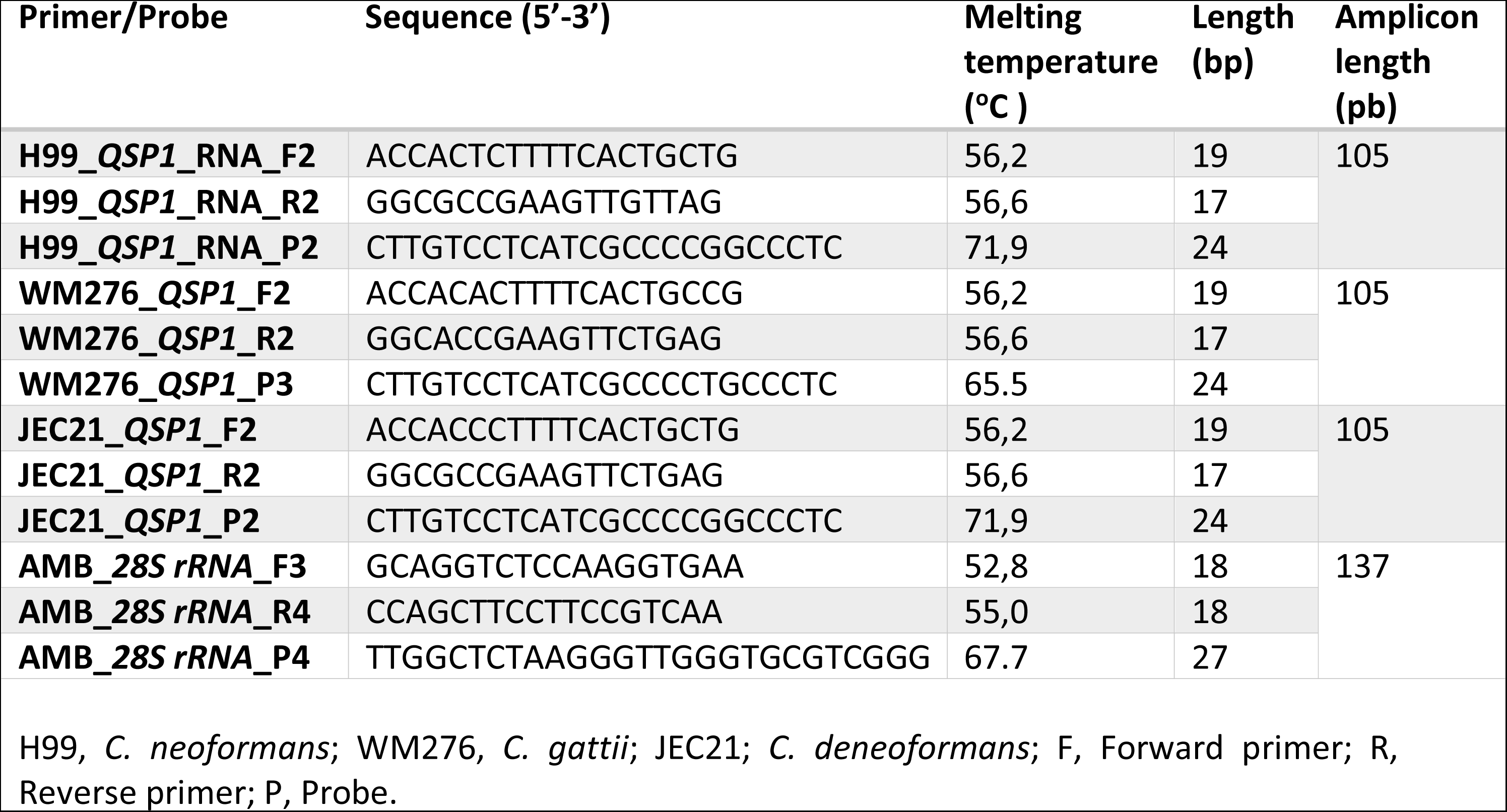
Species specific primer and probe sets that were designed and used in this study.

### Relative expression of QSP1A *in vitro*

A total of 10^6^ H99 cells were exposed 65°C for 2 hours (Heat killed) and 1mM of H_2_O_2_ (H_2_O_2_ killed) 1h with stationary phase as control and extracted using the same procedure as above. QSP1 mRNA expression was compared to actine (*ACT1)* mRNA expression using the method of Pfaffl, considering the efficacy of both assays (QSP1=2; ACT1=1.92) ^23^

### Quantification and qPCR efficiency

A 10-fold dilution from the strain H99 cells (*QSP1* A), WM276 cells (*QSP1* B/C) and JEC21 cells (*QSP1* D) cells were used to create a standard curve, and extrapolation was used to determine the absolute number of *Cryptococcus* cells present in the sample. Regression lines were constructed by plotting the logarithm of the template concentration versus the corresponding Cq value (Supplemental Figure 1A).

### Biosynex^®^ CryptoPS antigen screening in plasma

A semi-quantitative lateral flow *Cryptococcus* antigen (CrAg) test (Biosynex, Strasbourg, France) was performed following manufacturer’s instructions.^24,25^

### Study participants and samples and ethical approvals

We collected samples from 209 participants with CM from Botswana (85/205) and Malawi (124/205) enrolled in the AMBITION-cm randomised controlled trial.^3^ CSF samples collected at day 0 (D0), day 7 (D7) and day 14 (D14) post-antifungal treatment initiation were tested with QCC, and qPCR and RT-qPCR assays. Plasma samples collected at D0 were screened with a CryptoPS test. QCC and India Ink were performed as already described.^26^ All ethical approvals were described in previous work.^26,3^

### Statistical analysis

Graph Pad prism 9.4.0 (GraphPad, San Diego, California) was used for statistical analysis and data visualization. Kruskal-Wallis test was used to compare means between three or more matched groups when the data were not normally distributed, while Mann-Whitney tests were used for comparison of 2 groups of data when the data were not normally distributed. P-values <0.05 were considered statistically significant.

## Results

### Analytical specificity of *QSP1* (A, B/C, D) and *28S rRNA* assays on *Cryptococcus spp.* Strains

The *28S rRNA* assay was tested on a panel of 93 fungal species DNA, including species closely related to *Cryptococcus*, but also including Ascomycetes species such as *Candida* spp.*, Aspergillus spp., Saccharomyces spp., Fusarium spp., Penicillium spp*. (Supplementary Table 1). No cross reaction with non-*Cryptococcus* species complexes strains (100% analytical specificity) was observed. The *28S rRNA* assay amplified all *C. neoformans* and *C. gattii* species complexes with ratios between of 0.46-1.0 as compared to non-C. *neoformans/C. gattii* species complexes harbouring ratios between 0.0126 to 0.3322 (Supplementary Figure 1).

All *QSP1* assays (*QSP1* A, *QSP1*B/C and *QSP1* D) were tested on the same 93 fungal species and did not amplify *Filobasidium uniguttulatus*, *Vishniacozyma heimayensis* and *Salicoccozyma aerius* (Supplementary Figure 1)*. QSP1A* assay amplified serotype A DNAs with a ratio between 0.6113 and 1.00 except for VNII_T4, VNII_8 and VNBII_Bt1 where the amplification was lower with a ratio between 0.1224 and 0.2745. *QSP1A* assay was able to perfectly amplify VNIII (AD hybrid) with a ratio of 1, but not *C. deneoformans* and *C. gattii* species (Supplementary Figure 1). *QSP1*D assay amplified *C. deneoformans* and VNIII (AD hybrid) *with* a ratio between 0.673 to 1, but not *C. neoformans* and *C. gattii* species. *QSP1*B/C assay allowed amplification of *C. gattii* species, mainly on *C. gattii* s.s., *C. deuterogattii* and *C. tetragattii* (VGI VGIV, VGII) with ratio over 0.7195. However, *C. bacillisporus* DNAs were amplified with ratios < 0.1. *QSP1*B/C assay did not amplify *C. neoformans* and *C. deneoformans* DNA.

### Validation of the qPCR assays and extraction procedures

We first used spiked samples to optimize and validate our assays. The limit of detection with the *QSP1A* and the *28S rRNA* assay were 50 genomes and 1 genome per reaction, respectively. The efficiency of *QSP1A* assay was 1.98 (slope=-3.347, r^2^=0.998), while that of *28S rRNA* assay was 2.08 (slope=-3.141, r^2^=0.993, Supplemental Figure 2A). For comparison of pre-extraction procedures, amplification of untreated (control) condition after freezing gave a significantly lower Cq value than the other conditions including bead beating (BB), addition of proteinase K (PK) and BB+PK (p<0.0001, p=0.0297 and p<0.0001, respectively) (Supplementary Figure 2B), suggesting better extraction efficiency. There were no statistically significant differences between the two extraction protocols (Pathogen universal and DNA blood) of CSF tested on the MagNa Pure 96 instrument (Supplementary figure 2C).

### Prevalence of *Cryptococcus* species complexes in Botswana and Malawi

We designed species specific *QSP1* assays allowing rapid *Cryptococcus* identification in CSF specimens. We first validated *QSP1* identification with identification of isolates performed using whole genome sequencing. We obtained an adequate identification in 98.4% of the isolates (Appendix Table 2). Upon screening of clinical CSF samples from baseline (D0) in 209 participants, including 85 from Botswana and 124 from Malawi, we found a predominance of *C. neoformans*, 67% [95% CI: 55, 75] in Botswana and 68% [95% CI: 57, 73] in Malawi. The prevalence of *C. gattii* species complex was higher in Botswana (21% [95% CI: 14, 31]) than in Malawi (8% [95% CI: 4, 14]). A total of 8%, and 23% of the samples could not be serotyped in Botswana and Malawi respectively due to low fungal (*QSP1* negative, 28S positive). No pure *C. deneoformans* CM was detected in both countries. Of note, three (4%) and one (1%) samples were missing at D0 in Botswana and Malawi, respectively.

### Cryptococcal species-specific fungal load using *QSP1* and *28S rRNA* assays

We compared the quantification obtained in CSF pellet samples from *neoformans* and *gattii* CM using *QSP1* and *28S rRNA* qPCR assays based on Cq values. There was no statistically significant difference in the initial fungal loads (expressed as quantification cycles, Cq) of *C. neoformans* and *C. gattii* CM with *QSP1* assay (35.18 [30.45-38.36] vs. 34.57[31.19-37.32], respectively) or *28S rRNA* assay (28.77 [24.66-32.77] vs. 27.64 [24.87-31.73]) (Figure 2C). This was also observed for QCC, *QSP1* and *28S* assays quantification with absolute yeast number (Figure 3G-I). The difference in Cq value (ΔCq) between *QSP1* and *28S rRNA* assays for each CSF were significantly different with a ΔCq of 5.71 [IQR-5.09-6.32], reflecting about 50 copies of *28S rRNA* per genome in *C. neoformans* and 6.55 [5.75-7.78], reflecting about 100 copies of *28S rRNA* genome in *C. gattii*. This has also been observed in the DNA extracted from strains during prior optimizations (Supplemental Figure 2D).

**Figure 2:**
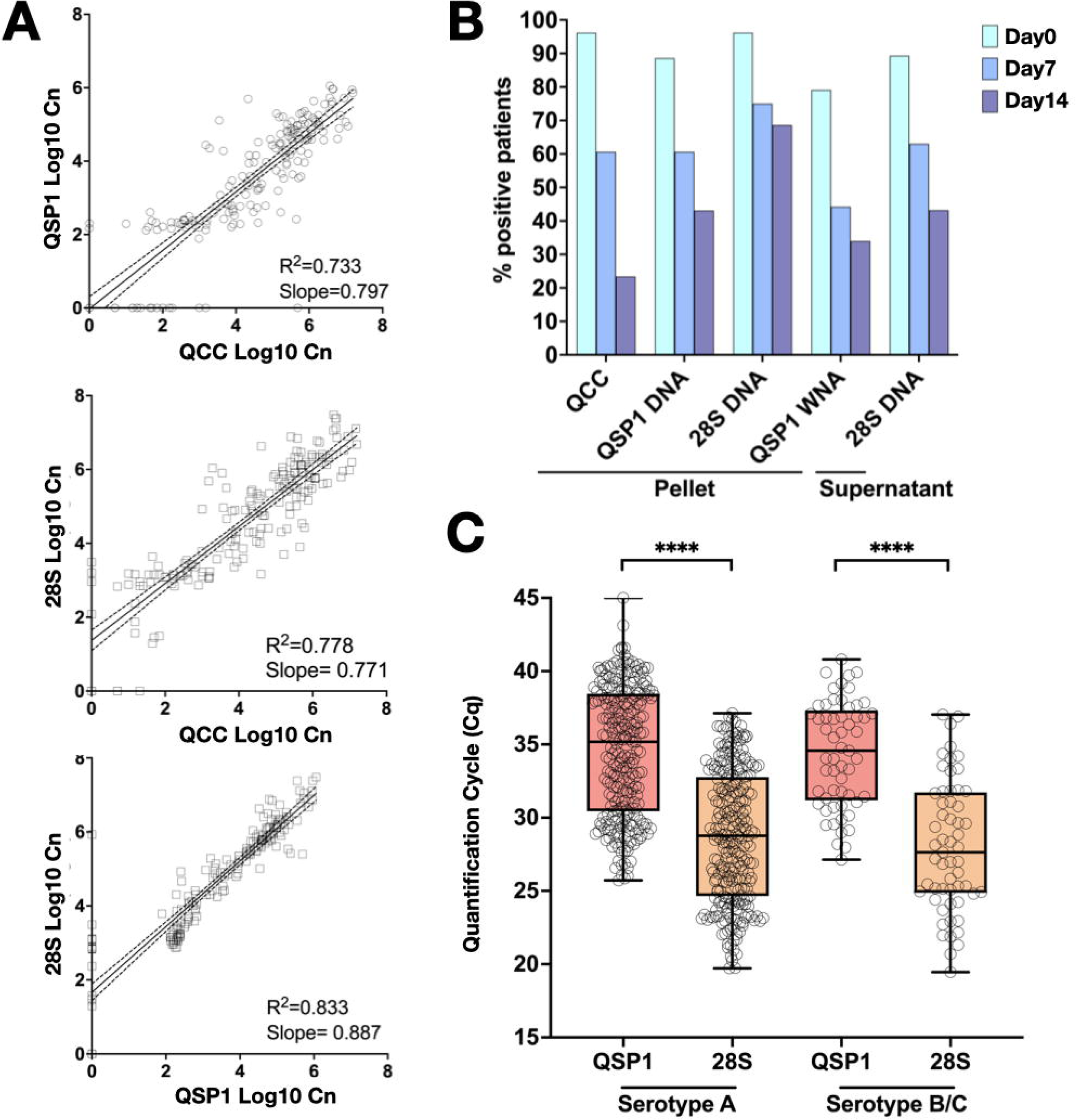
Positive correlations between *QSP1* quantification (Log_10_ Cn cells/mL, QSP1 Log10 Cn) and QCC (Log_10_ Cn cells/mL, QCC Log10 Cn), *28S rRNA* quantification (Log_10_ Cn cells/mL, 28S Log10 Cn) and QCC and *28S rRNA* and *QSP1* using CSF pellet results at day 0 (D0, before starting antifungal treatment) (A). Percentage of CSF pellet and supernatant samples amplified by *QSP1* A, B/C and *28S rRNA* at Day 0 (D0), Day 7 (D7) and Day 14 (D14) of antifungal treatment (B). Fungal loads in CSF at D0 with *QSP1* and *28S rRNA* assays expressed as quantification cycles in *C. neoformans* and *C. gattii* (C). ****, p<0.0001; ns, not significant.

**Figure 3:**
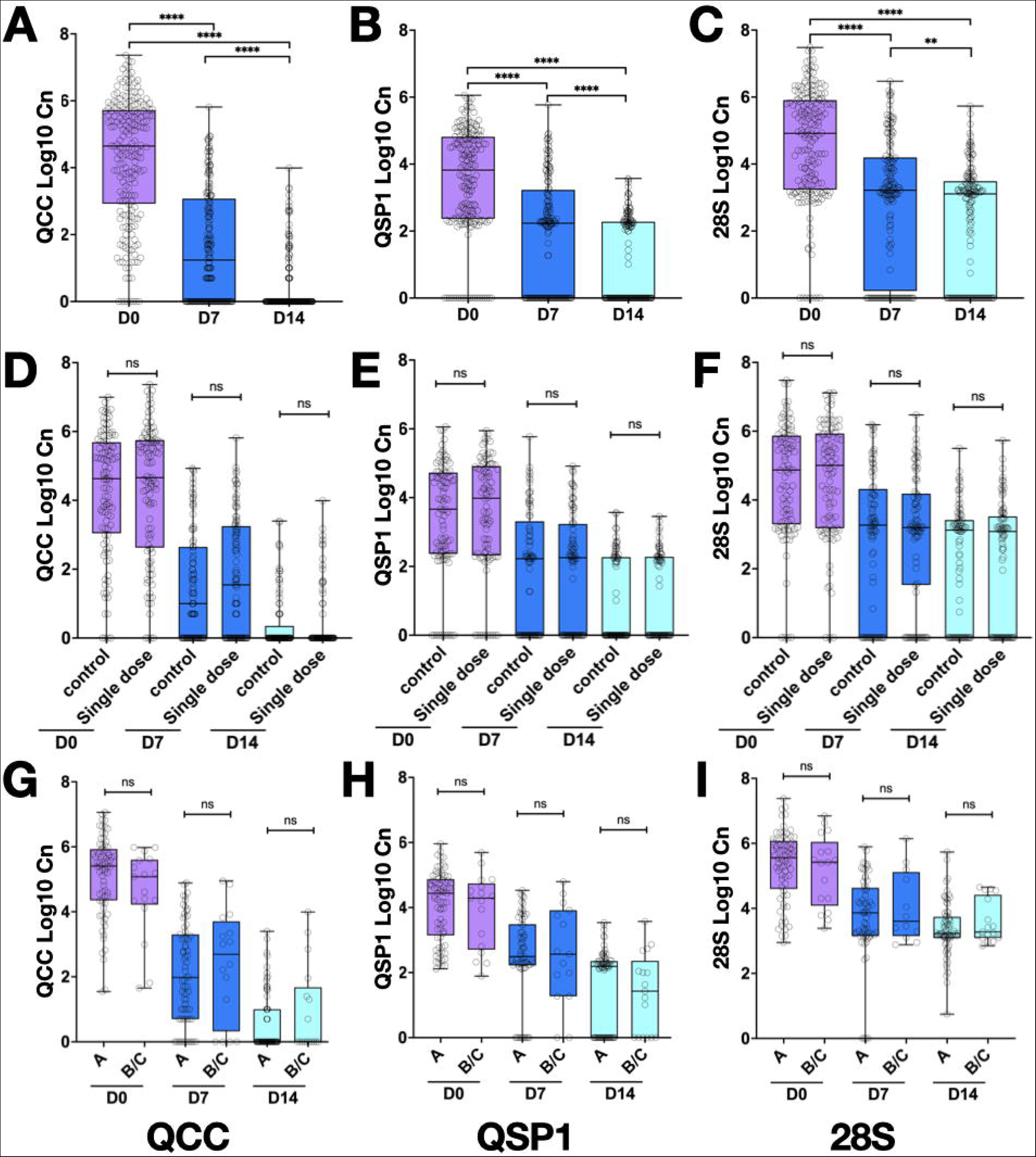
Cryptococcal fungal load quantification (Log_10_ Cn cells/mL, Log10 Cn) at Day 0 (D0), Day 7 (D7) and Day 14 (D14) in CSF of participants by QCC, *QSP1* assay, and *28S rRNA* assay in total populations (A,B,C respectively) and in the control or single dose regimens (D, E, F respectively). Serotype specific fungal load quantification at Day 0 by QCC, *QSP1* and *28S rRNA* (G,H,I). **, p<0.001 ****, p<0.0001; ns, not significant.

### Comparison of quantification using our qPCR assays on different fractions of CSF samples and QCC

We quantified DNA using *QSP1* assays and *28S* assays in CSF pellets and in CSF supernatant. We also quantified WNA (RNA+DNA) in the CSF pellet as a potential proxy of the viability of the yeasts present in the pellet. At D0 in CSF pellets, the proportion of positivity with DNA was higher with QCC and *28S rRNA*, with 95% and 94% of positive participants, respectively as compared to *QSP1*, where 88% were positive (Figure 2B). At D14, *QSP1* and *28S rRNA* assays showed an increased proportion of positive samples (43.1% and 68.6%, respectively) as compared to QCC (23.4%).

At D0 in CSF pellets, the sensitivity of the *28S rRNA* and *QSP1* assays were 98.2% [IC95 95.1-99.5] and 90.4% [IC95 85.2-94.0] when compared to QCC as the reference standard. Indeed, three QCC-negative CSF were found to be *28S rRNA*-and *QSP1*-positive. The sensitivity was 98.0 [IC95 94.4-99.5] and 93.0% [IC95 87.9.1-96.0] when compared to the India ink results. Of note, 27 and 16 India ink-negative CSF samples were found to be positive with *28S rRNA* and *QSP1*, respectively. Specificity could not be calculated as all CSF samples came from participants with CM.

Detection of *28S rRNA* DNA in CSF supernatant samples gave a lower proportion of positivity when compared to CSF pellet samples with a maximum difference at D14 (43.2 vs. 68.6%) (Figure 2B). *QSP1* WNA detection positivity was lower as compared to *QSP1* DNA in CSF pellets samples at all timepoints (Figure 2B) but was still more positive at D14 than in samples that were quantified with QCC (34 vs. 23.4% positivity). A Bland-Altman analysis shows an agreement between QCC and *QSP1* or 28S with a ratio bias of 1.24±0.28 and 0.90 ± 0.24, respectively (Supplemental Figure 3)

We compared the quantification of fungal load with QCC, *QSP1* and *28S rRNA* qPCR assays in CSF pellets samples at D0. We found strong correlations between *QSP1*, *28S rRNA* and the gold standard QCC assay (Figure 2A), with a positive linear relationship between *QSP1* and QCC, R^2^=0.733 (Figure 2A), *28S rRNA* and QCC, R^2^=0.778 (Figure 2A), and the R^2^ value between the designed assays, *QSP1* vs *28S rRNA* was 0.833 (Figure 2A).

### Significant decrease of fungal load between D0 and D14 upon treatment initiation with QCC, *QSP1* and *28S rRNA* but no cryptococcal fungal load differences between treatment arms

*QSP1* and *28S rRNA* assays showed a significant decrease in cryptococcal fungal load between D0 and D7, D7 and D14, and D0 and D14 (p<0.0001) (Figure 3B, 3C), which is similar to the decrease observed with QCC (Figure 3A) . Of note, fungal load dynamics was similar in patients from each arm, thereby suggesting a similar fungicidal activity of the two strategies (Figure 3 D, 3E, 3F).

### Comparison of CrAg detection in plasma at D0 and quantification of fungal load with QCC, *QSP1* and *28S rRNA* in initial CSF

The 151 participants plasma samples which were determined as serotype A or B/C, were tested with CryptoPS at D0, 98 (64.5%) had a positive T1 & T2 band, 49 (32.5%) a positive T1 band and 4 (2.6%) were negative. Negative CryptoPS tests were identified in serotype B/C infections (4/24, 16.7%), while all serotype A participants had a positive CryptoPS test in the plasma at D0. This was validated with WGS identification as all negative CryptoPS tests came from *C. tetragattii* cases (Appendix Table 2). In serotype B/C, 4 and 3 samples were negative with CrAg while positive with QCC (Figure 4A) and *QSP1* (Figure 4B) and *28S rRNA* (Figure 4C), respectively (data was missing for one sample using *28S rRNA* assay).

**Figure 4:**
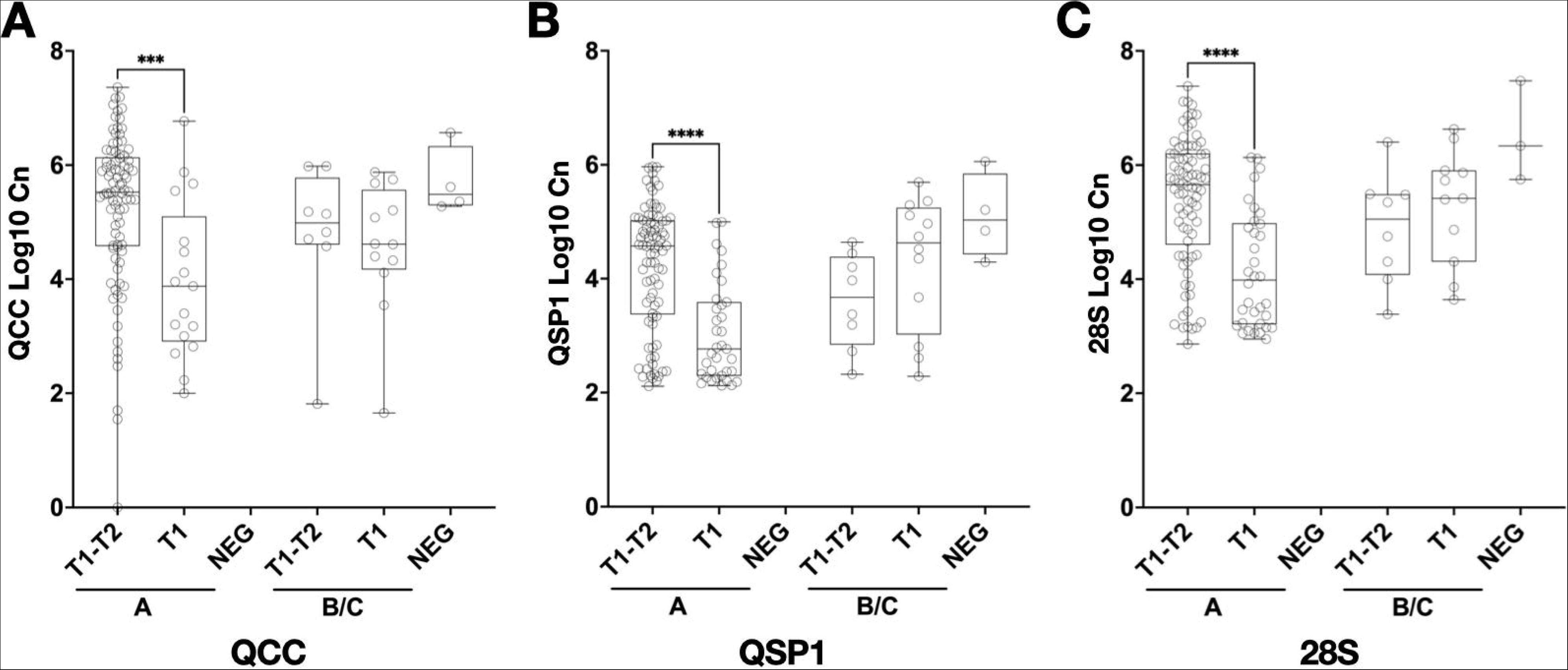
Quantification of the fungal load in CSF (Log_10_ Cn cells/mL, Log10 Cn) of participants with T1-T2 positive band, T1 band and negative CryptoP/S testing in plasma at Day 0 (D0) in serotype A or B/C infections with QCC (A), *QSP1* (B) and 28S (C) qPCR. ***, p<0.001; ****, p<0.0001

### Quantification of yeasts cells in CSF of participants are mainly from viable and culturable cells but can also be viable but non culturable cells (VBNC)

We also wanted to determine if the DNA detected and quantified in CSF came from viable yeasts. We first validated that dead cells *in vitro* had a decreased expression of QSP1 as compared to the actine gene (*ACT1*) and to stationary phase *Cryptococcus* cell (Supplemental Figure 2F).

We used the same *QSP1A* assay but with a reverse transcriptase quantitative PCR (RT-qPCR) allowing the detection and quantification of both *QSP1* mRNA and DNA (WNA amplification). We found that there was an increased quantification of the *QSP1A* using RT-qPCR at all timepoints (D0, D7, D14), p<0.0001, Figure 5 A-C). A gain between 0.8 (D0) and 1.1 (D7 and D14) Log Cn cells was observed representing 10-fold more detection of WNA than DNA. This proved that RNA target was present in a higher quantity (around 10 times more) than the *QSP1* DNA gene. During optimizations using the H99 strain, we also found an increased quantification with the RT-qPCR as compared to the qPCR with quantification cycle values of 26.55 [22.86-32.59] and 30.61 [27.46-36.61], p<0.0001, respectively (Supplemental Figure 1E).This suggested that yeasts cells detected in the CSF pellet of the participants were viable, producing more *QSP1* mRNA target than DNA target present in the eluate, showing an active expression of *QSP1* gene in a viable organism.

**Figure 5:**
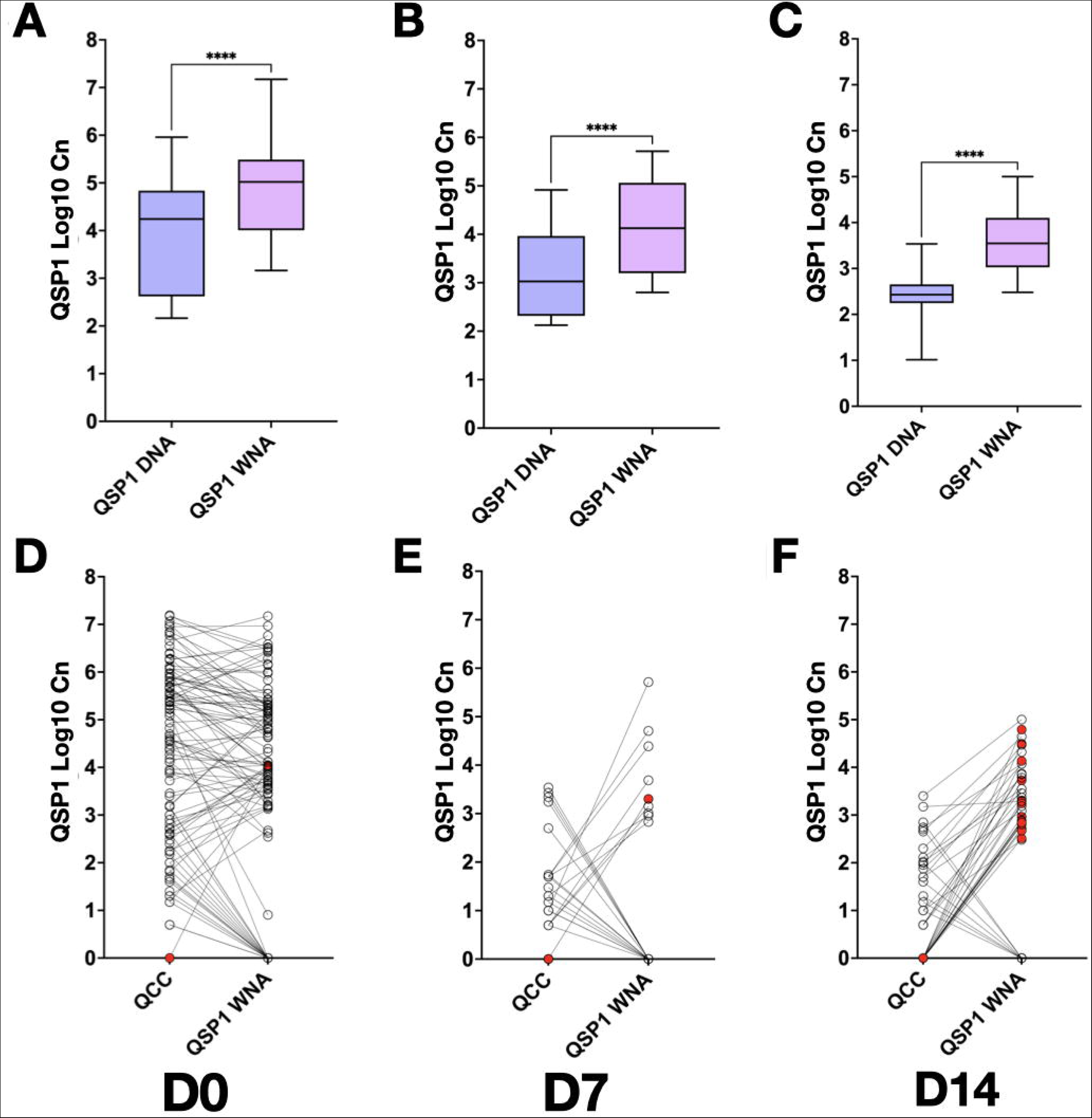
*QSP1* DNA (qPCR) and WNA (RTqPCR) quantification in CSF pellet at Day 0 (D0) (A), Day 7 (D7) (B) and Day 14 (D14) (C) using *QSP1* assays (A-C), and comparison between QCC and *QSP1* WNA load at Day 0 (D0), Day 7 (D7) (E) and Day 14 (D14) (F). ****, p<0.0001. Red circles are participants with negative QCC and positive *QSP1* WNA detection

We then compared QCC load and *QSP1* WNA load in individual samples. We noticed that QCC was negative and *QSP1* WNA positive in one sample at D0 and D7 and 17 samples at D14 (Figure 5 D-F, red circles). We thus individualized patients with a negative CSF culture but a positive *QSP1* RT-qPCR assay suggesting that viable but non culturable cryptococci were observed in the CSF during meningitis. The W10 outcome of these patients was 23.5%.

## Discussion

Several qPCR and RT-qPCR assays (*QSP1A*, *QSP1B/C* and *QSP1D* and *28S rRNA* assays) allowing *Cryptococcus* load quantification and specific identification of *C. neoformans*, *C. deneoformans and C. gattii* species complexes were designed, optimized, and clinically evaluated using samples collected during the AMBITION-cm trial.^3^ The analytical specificity and the sensitivity have been validated and show optimal performance, following dedicated MIQE guidelines.^10^ We report optimal qPCR efficiencies of ∼100% for all assays. *QSP1* and *28S rRNA* assays were able to detect specified species, as shown in Supplementary figure 2A. Based on our results, we therefore recommend using *28S rRNA* assay as a screening assay and when positive to identify the species and quantify the fungal load with *QSP1* species specific assays.

Interestingly, we identified a high burden of *C. gattii* species complex infections in Botswana (21%) as compared to Malawi (8%) as already reported in those countries (being, 13% and 30% for Malawi and Botswana, respectively).^27,28^ No *C. deneoformans* (serotype D) cases was detected in the cohort, as already shown in patients native from Africa and leaving in Europe.^29^

The designed *QSP1* and *28S rRNA* assays showed excellent correlation with QCC quantification. QCC is the current gold standard in cryptococcal fungal load quantification with a reported 94.2% sensitivity in South Africa and Uganda.^13^ We clearly observed a gap between QCC and DNA detection with an increased number of participants detected with the *QSP1* and *28S rRNA* qPCR assays than QCC, suggesting that yeasts unable to grow on culture media or dead yeasts can be detected by qPCR especially at low fungal loads. Nevertheless, participants had lower fungal loads at D7 and/or D14, than the initial load at D0, demonstrating the early fungicidal activity of the two different treatment arms. This finding is important as a decrease of the fungal load and a fungicidal activity could be determined using qPCR quantification instead of QCC.^17^ We are currently working at validating our assay in patients with CM and receiving both antiretroviral and antifungal therapies to discriminate relapse from immune reconstitution inflammatory syndrome.

By using whole nucleic acids amplification with our *QSP1* assay, which allows amplification of *QSP1* mRNA and DNA, we validated here that most of the yeasts detected, even at D14, were viable, with an increased WNA detection compared to DNA detected. We thus detected DNA (qPCR) in participants with negative QCC results at D0 D7 and D14 and significantly more amplification with *QSP1* WNA (RT-qPCR), suggesting that living yeasts were present in the CSF despite a negative culture at D0, D7 or D14. These cells are suggestive of viable but non culturable cells (VBNC), meaning viable living yeasts but not capable of growth on agar plates. This phenotype is known in *Cryptococcus* yeasts cells and have been identified and characterized recently by our team.^30,31^ This is a major finding as RT-qPCR should be better at characterizing the presence of viable cells as compared to QCC. It tends to show that VBNC can be found in CSF and that antifungal treatment could induce this phenomenon. More work needs to be done to investigate those VBNC in human CSF under treatment.

In a study done by Tenforde et al. in Botswana assessing the sensitivity and specificity of CryptoPS, the sensitivity was found to be 61.0% and the specificity to be 96.6%.^24^ Additionally, it has been shown recently that the CryptoPS test was not detecting in vitro capsular antigens from *C. gattii* species complex including *C. bacillisporus*, *C. deuterogattii* and *C. tetragattii* species.^32^ Our results show that there are false negative results only in serotype B/C infections but not with serotype A. The high prevalence of serotype B/C (*C. gattii* species complex) infections in Botswana (see above) might explain the low sensitivity rate that was previously reported when using CryptoPS locally.

In conclusion, we designed and validated three qPCR in three assays (*QSP1* A, *QSP1* B/C, and *28S rRNA*) for the detection and quantification of *Cryptococcus* spp. (*28S*) in CSF. These assays have excellent correlation with the current gold standard, i.e QCC. qPCR will thus provide easier fungal load monitoring tool and early information on subsequent outcome during HIV-associated cryptococcal meningitis in sub-Saharan Africa. As a diagnostic tool that can speciate and give fungal load, it enables stratified management of patients in the future. Our assays can be used as research tools to measure clearance more quickly and easily as an endpoint to assess novel antifungal regimens.

## Data Availability

All Data will be available upon request to

## Acknowledgments

We thank Vincent Enouf and Tiffany Dos Santos from the P2M facility, Institut Pasteur for their help on the MagNa Pure instrument. We warmly want to thank Philippa Griffin-Johnstone, the administrative manager during the Ambition trial her extensive and precious administrative support. We would also like to thank the AMBITION-cm team, especially personnel in Blantyre and Gaborone, together with Dr Sumayah Salie, Dr Charlotte Schutz and Dr Muki Shey in UCT for their assistance with paperwork in UCT. We thank the AMBITION-cm trial participants and their families and caregivers, as well as all the clinical, laboratory, and administrative staff at all the sites who were not directly involved in the trial; Andrew Nunn, Sayoki Mfinanga, Robert Peck, and William Powderly for serving on the data and safety monitoring committee; and John Perfect, Andrew Kambugu, Saidi Kapigi, and Douglas Wilson for serving on the trial steering committee.

## The AMBITION study group

In addition to the named authors, the following were members of the Ambition Study Group:

Botswana Harvard AIDS Institute Partnership / Princess Marina Hospital, Gaborone, Botswana – J Goodall, N Mawoko, J Milburn, R Mmipi, C Muthoga, P Ponatshego, I Rulaganyang, K Seatla, N Tlhako and K Tsholo.

University of Cape Town / Mitchells Plain Hospital / Khayelitsha District Hospital, Cape Town, South Africa – S April, A Bekiswa, L Boloko, H Bookholane, T Crede, L Davids, R Goliath, S Hlungulu, R Hoffman, H Kyepa, N Masina, D Maughan, T Mnguni, S Moosa, T Morar, M Mpalali, J Naude, I Oliphant, S Sayed, L Sebesho, M Shey and L Swanepoel.

Malawi-Liverpool-Wellcome Trust Clinical Research Programme / Queen Elizabeth Central Hospital, Blantyre, Malawi – M Chasweka, W Chimang’anga, T Chimphambano, E Dziwani, E Gondwe, A Kadzilimbile, S Kateta, E Kossam, C Kukacha, B Lipenga, J Ndaferankhande, M Ndalama, R Shah, A Singini, K Stott and A Zambasa.

UNC Project, Kamuzu Central Hospital, Lilongwe, Malawi – T Banda, T Chikaonda, G Chitulo, L Chiwoko, N Chome, M Gwin, T Kachitosi, B Kamanga, M Kazembe, E Kumwenda, M Kumwenda, C Maya, W Mhango, C Mphande, L Msumba, T Munthali, D Ngoma, S Nicholas, L Simwinga, A Stambuli, G Tegha and J Zambezi.

Infectious Diseases Institute / Kiruddu General Hospital, Kampala, Uganda – C Ahimbisibwe, A Akampurira, A Alice, F Cresswell, J Gakuru, D Kiiza, J Kisembo, R Kwizera, F Kugonza, E Laker, T Luggya, A Lule, A Musubire, R Muyise, O Namujju, J Ndyetukira, L Nsangi, M Okirwoth, A Sadiq, K Tadeo, A Tukundane and D Williams.

Infectious Diseases Institute / Mbarara Regional Referral Hospital, Mbarara, Uganda – L Atwine, P Buzaare, M Collins, N Emily, C Inyakuwa, S Kariisa, J Mwesigye, S Niwamanya, A Rodgers, J Rukundo, I Rwomushana, M Ssemusu and G Stead.

University of Zimbabwe / Parirenyatwa General Hospital, Harare, Zimbabwe – K Boyd, S Gondo, P Kufa, E Makaha, C Moyo, T Mtisi, S Mudzingwa, T Mwarumba and T Zinyandu.

Institut Pasteur, Paris, France - Françoise Dromer

London School of Hygiene and Tropical Medicine, London, UK – P Griffin and S Hafeez.

## Figure Legend

**Supplementary figure 1: Analytical specificity of the different assays.** A heat map showing amplification of *C. neoformans*, *C. gattii* species and other *Cryptococcus* species (*F.uniguttulatus*, *V. haemaeyensis*, *S. aerius*) by *28S rRNA*, *QSP1A*, *QSP1*D and *QSP1*B/C assays based on the ratio of Cq value tested / Cq value of the reference DNA.

*Serotype A (VNI, VNII, VNBI, VNBII), serotype D (VNIV), serotype B/C (VGI, VGII, VGIIIa, VGIIIb, VGIIIc,VGIV).

Serotype AD (VNIII). Ratios 0-0.3 indicate no amplification to poor amplification, 0.4-0.6 low amplification, and 0.7-1.0 good to very good amplification.

**Supplementary Figure 2: Implementation and technical validation of the different qPCR assays.** Efficiency of *QSP1* A and *28S rRNA* (A), untreated condition yielded more DNA than treatment with bead beating (P<0.001) and proteinase K (p=0.0297)(B). There were no differences in nucleic acid extractions between pathogen universal and DNA blood protocol (C). *QSP1*A and *28S rRNA* Cq from simulated CSF samples seeded with different concentrations of H99 (D) and comparison of Cq between *QSP1*A DNA and *QSP1*A WNA using H99 cells (E). ns, not significant; *, p<0.05; ****, p<0.0001. Relative expression of *QSP1* mRNA to actine (ACT1) mRNA in heat killed or H_2_O_2_ killed H99 Cn cells as compared to stationary phase H99 (F).

**Supplementary Figure 3: Blant-Altman analysis of** Bland-Altman analysis comparing cell quantification between QCC and *QSP1*A (A) or 28S (B) by plotting the ratio of each pair of value against the average of them. The lines represent the bias (solid line) and the 95% agreements (dotted lines).

**Supplementary table 1:**
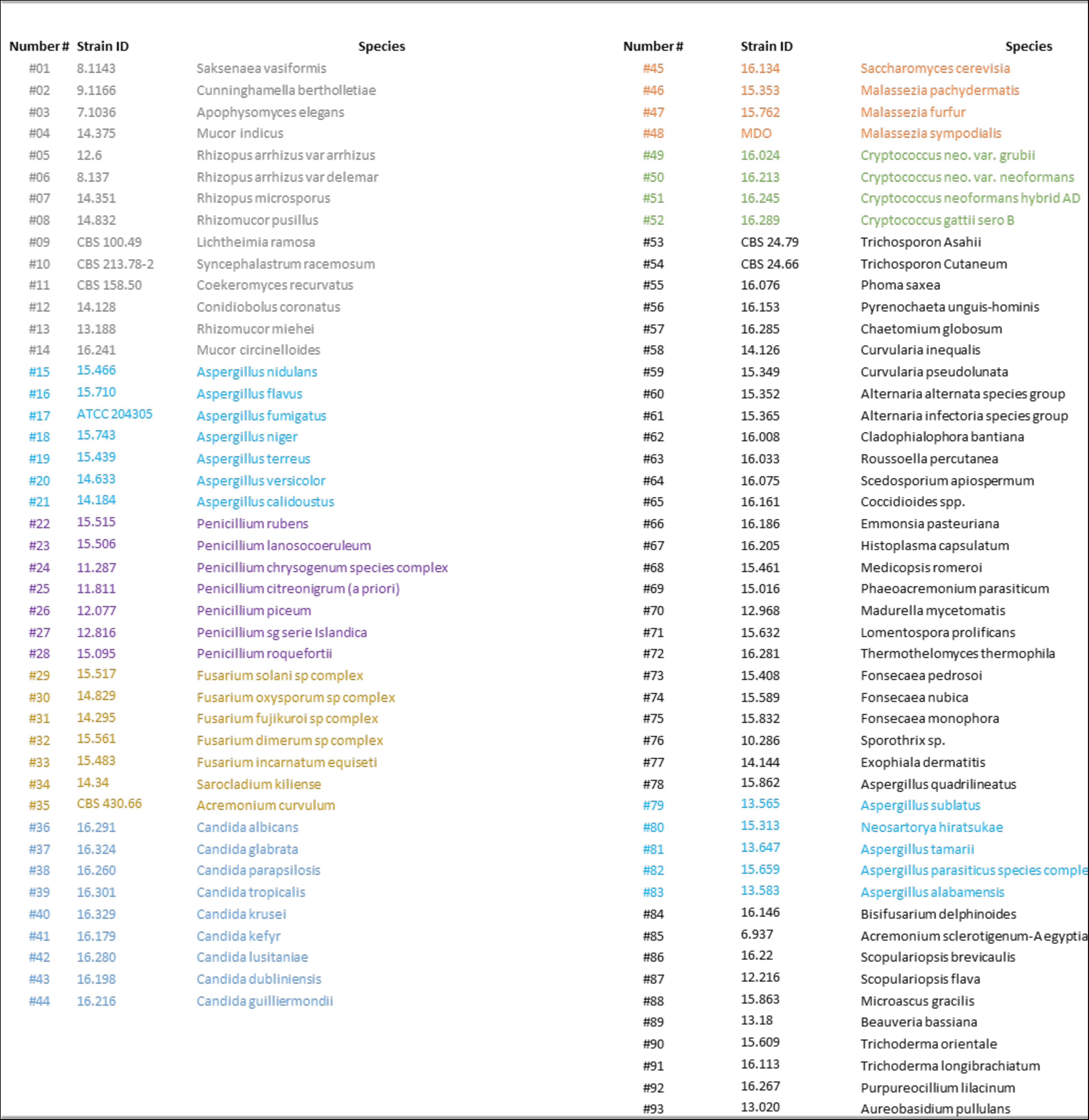
Fungal species DNA used to test the analytical specificity of the primers.

**Supplementary table 2 :** Correlation between whole genome sequencing identification of the recovered isolate and *QSP1*A identification and CrAg testing (Biosynex test) in CSF and plasma.

|  | qPCR assay |  |  | Biosynex in CSF |  |
| --- | --- | --- | --- | --- | --- |
|  | *QSP1A* | *QSP1B/C* | *QSP1D* | Negative | Positive |
| *Cryptococcus* species upon WGS of isolate | n (%) | n (%) | n (%) | n (%) | n (%) |
| *Cryptococcus neoformans* | **104/104 (100)** | ND | ND | 0/101 (0) | **101/101 (100)** |
| *C. neoformans C. deneoformans* hybrids | **4/4 (100)** | ND | ND | 0/4 (0) | **4/4 (100)** |
| *Cryptococcus tetragattii* | 2/16 (12.5) | **14/16 (87.5)** | ND | 3/13 (23) | **10/13 (77)** |
| *Cryptococcus gattii* | 0 (0) | **4/4 (100)** | ND | 0/4 (0) | **4/4 (100)** |
| ND, not done |  |  |  |  |  |

