## Supplementary figures and images for "Innovative quantitative PCR assays for the assessment of HIV-associated cryptococcal meningoencephalitis in Sub-Saharan Africa"

### Supplemental Figure 1

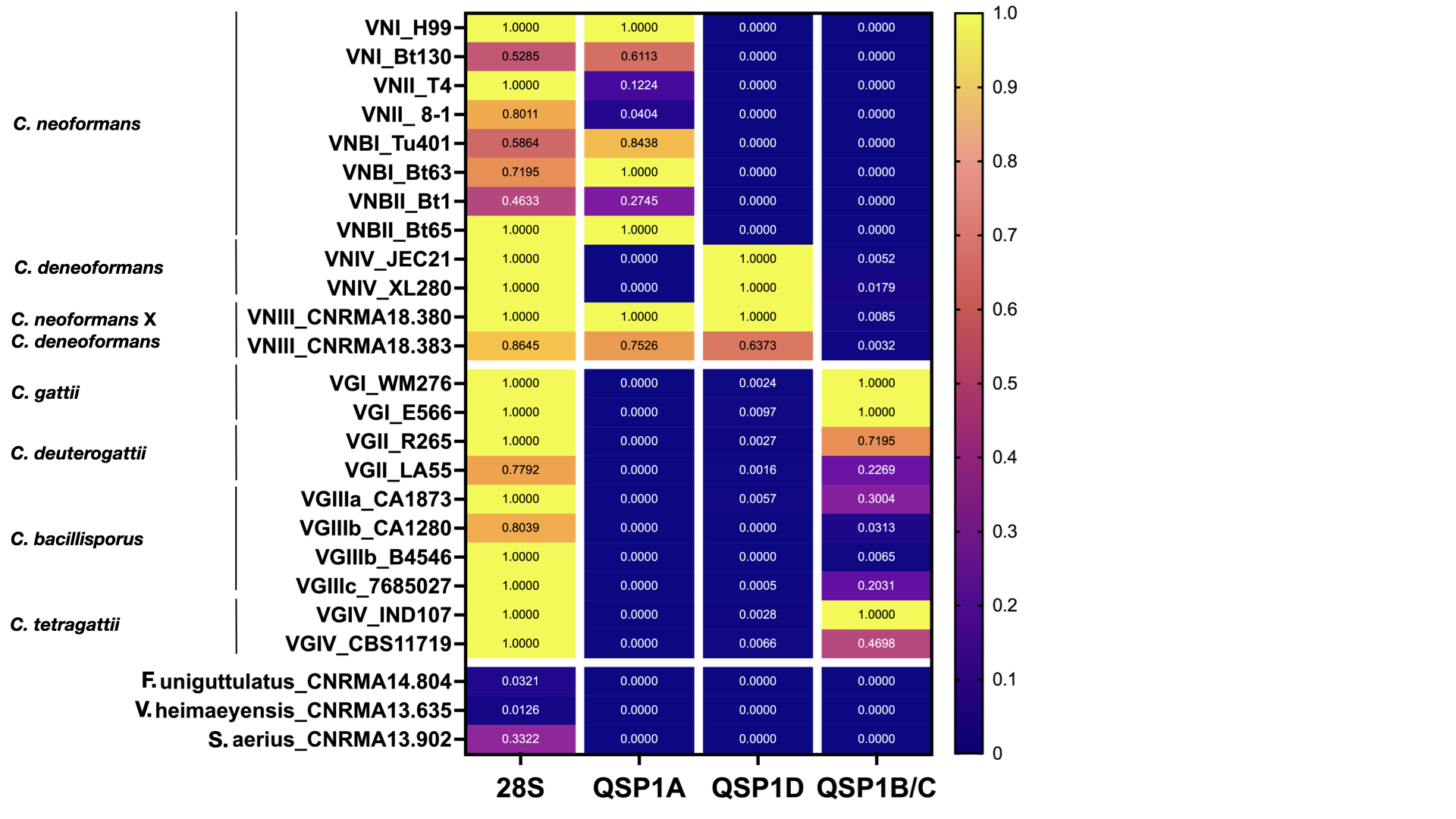

### Supplemental Figure 2

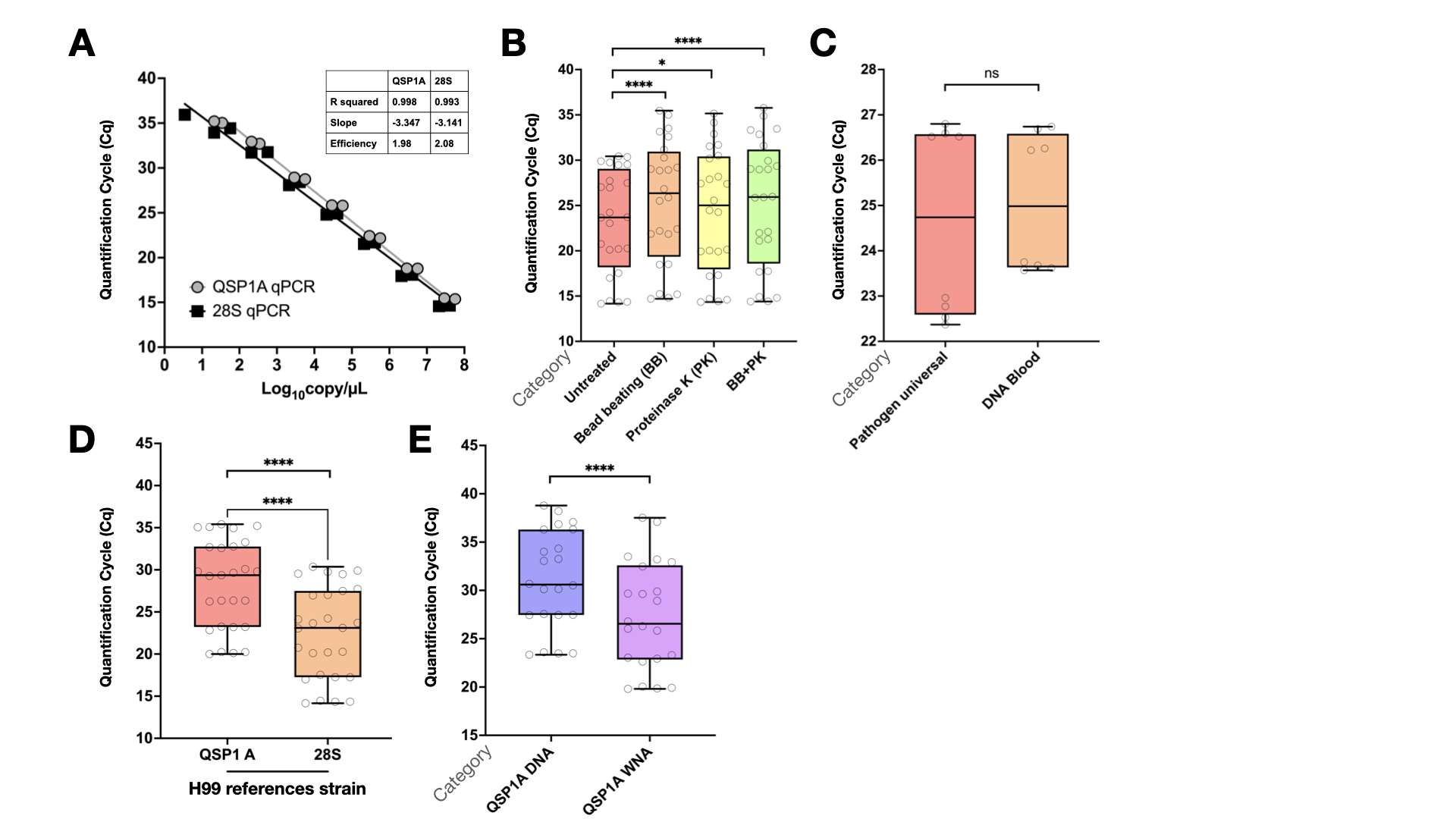

### Supplemental Figure 3

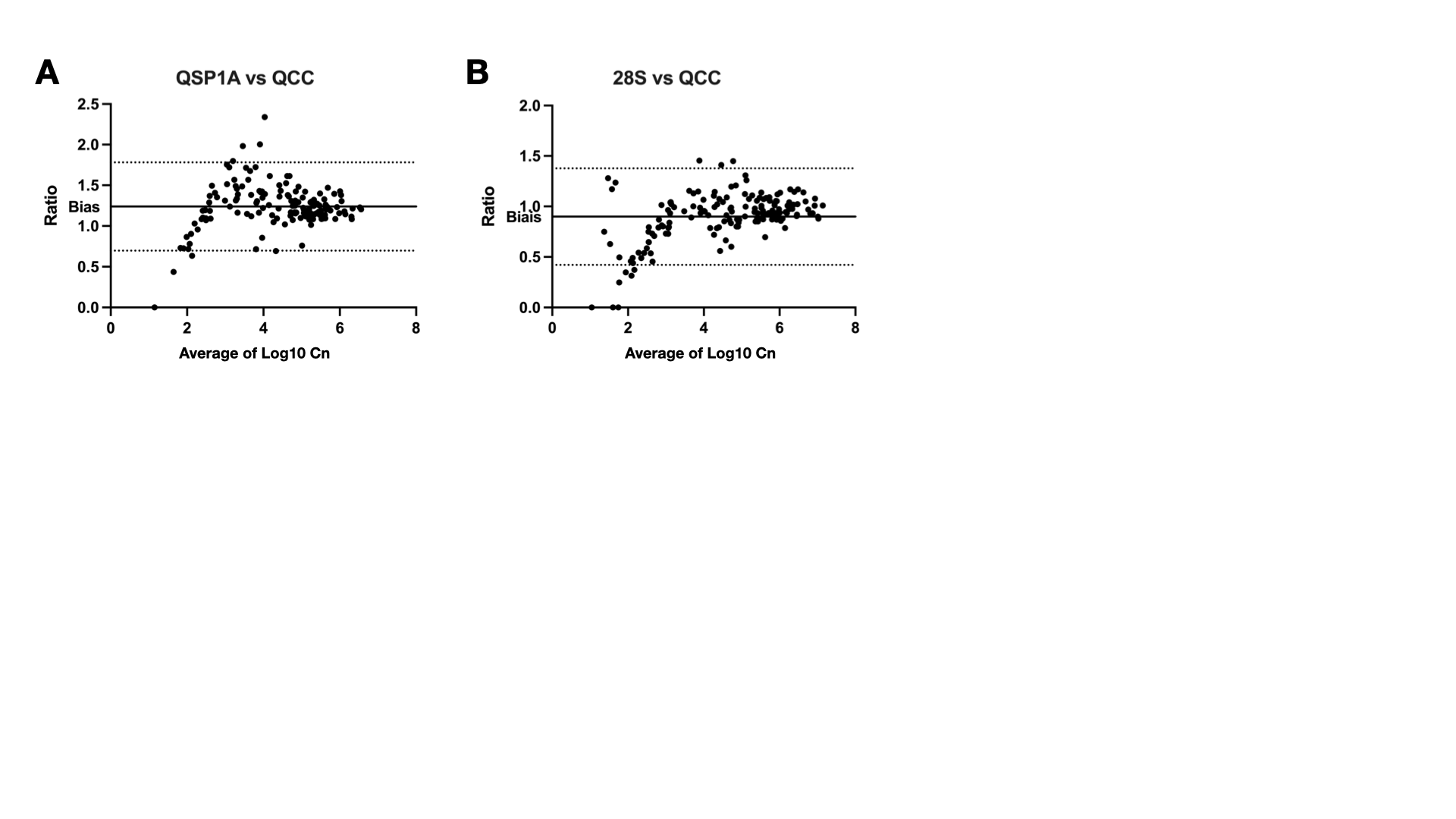
